# Post-COVID Spirometric Abnormalities in Workers with Intermittent High-Altitude Exposure: A Cross-Sectional Study in Peru

**DOI:** 10.1101/2025.08.01.25332622

**Authors:** Jair Alonso Góngora-Bendezú, Marleyssi Valeria Martinez-López, Angel David Aguinaga-Fernandez, Marlon Yovera-Aldana

**Affiliations:** Maestría en Medicina Ocupacional y Medio Ambiente, Universidad Científica del Sur, Lima, Perú; Grupo de investigación de Neurociencias, Metabolismo, Efectividad Clínica y Sanitaria, Universidad Científica del Sur, Lima, Perú

**Keywords:** Spirometry, COVID-19, occupational exposure, altitude, pulmonary function

## Abstract

**Introduction:** SARS-CoV-2 infection may lead to persistent pulmonary sequelae, particularly relevant among workers exposed to occupational stressors such as high altitude. This study aimed to assess the prevalence and associated factors of spirometric alterations in workers with a history of COVID-19 and intermittent exposure to high altitude.

**Methods:** A cross-sectional analytical study was conducted in 400 sea-level-born workers intermittently exposed to altitudes above 2500 meters above sea level, all with a confirmed history of COVID-19. Only spirometry tests rated as quality A or B performed in 2024 were included. Sociodemographic, clinical, and occupational variables were evaluated. Adjusted prevalence ratios (aPR) were estimated using Poisson regression with robust variance.

**Results:** A total of 72.2% of workers showed spirometric abnormalities: 40.5% had a mixed pattern, 20.8% restrictive, and 11.0% obstructive. Independent factors associated with altered spirometry included obesity (aPR: 1.35; 95% CI: 1.19–1.53), high Charlson comorbidity index (aPR: 1.49; 95% CI: 1.33–1.68), prior occupational exposure to harmful substances for over 5 years (aPR: 1.64; 95% CI: 1.43–1.89), intermittent high-altitude exposure ≥7 years (aPR: 1.81; 95% CI: 1.60–2.05), and severe COVID-19 (aPR: 1.65; 95% CI: 1.41–1.91).

**Conclusions:** Over 70% of participants showed spirometric impairment was found in post-COVID-19 workers with intermittent high-altitude exposure. Respiratory function monitoring should be reinforced in this occupational group, especially among those with higher clinical and environmental risk factors.

## Introduction

Coronavirus disease 2019 (COVID-19), caused by the SARS-CoV-2 virus, has resulted in a substantial burden of long-term sequelae, particularly involving the respiratory system. Even in individuals without prior pulmonary conditions, post-infectious abnormalities in lung function have been frequently reported months after recovery, with clinical presentations ranging from persistent dyspnea to subclinical changes detectable only via pulmonary function testing [1]. Among these abnormalities, restrictive, obstructive, and mixed ventilatory defects have been observed, reflecting diverse underlying pathophysiological mechanisms such as interstitial inflammation, airway remodeling, and microvascular damage[2,3].

While these sequelae are increasingly documented in hospital-based and community populations, little is known about their impact in occupational groups exposed to extreme environmental conditions such as high-altitude workers.[4] Chronic or intermittent exposure to hypobaric hypoxia induces physiological adaptations, including increased pulmonary artery pressure, polycythemia, and vascular remodeling, but may also predispose susceptible individuals to maladaptive responses and impaired pulmonary recovery after infections[5]. Moreover, miners and other laborers at altitude frequently encounter additional hazards, including exposure to particulate matter, diesel fumes, and welding gases, all of which have been implicated in chronic airway inflammation and small airway disease[6,7]

In occupational health surveillance programs in Peru and other Andean countries, spirometry is commonly used as part of functional evaluations to assess respiratory fitness for high-altitude labor.[8] However, the COVID-19 pandemic disrupted baseline assessments and introduced a new, poorly characterized risk factor for post-infectious pulmonary impairment.[9] Importantly, the combined effects of SARS-CoV-2 infection, metabolic risk factors such as obesity, and environmental exposures have yet to be evaluated in a population with documented normal lung function prior to the pandemic.[10]

Recent evidence suggests that obesity, advanced age, and pre-existing comorbidities increase the likelihood of persistent pulmonary dysfunction post-COVID-19, even in mild cases[11]. However, few studies have explored the long-term respiratory impact of COVID-19 in high-altitude workers with pre-pandemic normal spirometry, where hypoxia and inhaled irritants may exacerbate residual inflammation or hinder tissue repair.[12]

Although some longitudinal studies have explored the respiratory sequelae of COVID-19 in clinical populations, there is a lack of evidence regarding post-COVID spirometric alterations in occupational settings—particularly among workers with intermittent high-altitude exposure, which is already an additional environmental risk on top of other potential respiratory risk factors present in their specific job activities. This study aims to address this gap by evaluating the prevalence and associated factors of abnormal spirometry patterns in this specific working population.

## MATERIALS AND METHODS

### Study Design and Setting

We conducted a cross-sectional analytical study using data from occupational medical evaluations conducted in 2024 at a high-altitude occupational health center located in Cusco, Peru, at an elevation of 3,400 meters above sea level. The facility provides routine medical evaluations for workers from four economic sectors: mining, administration, construction, and the preservation industry.

### Population and Sampling

The study population included male and female workers residing at sea level who had a confirmed diagnosis of coronavirus disease 2019 (COVID-19) by reverse transcription polymerase chain reaction (RT-PCR) between 2020 and 2021 and underwent comprehensive annual occupational medical evaluations in 2024, including spirometry. Only spirometric tests graded as A or B in quality, according to the standards of the American Thoracic Society and European Respiratory Society (ATS/ERS), were included.[13] Participants were required to have no prior history of chronic pulmonary disease and to have had normal spirometric values before the onset of the COVID-19 pandemic.

We excluded workers currently performing tasks with high occupational risk for chronic respiratory conditions, such as those exposed to silica, asbestos, mineral dust, or industrial vapors (e.g., textile workers processing pita fiber, miners, and construction workers exposed to cement or inorganic particulates). Workers with incomplete or inconsistent medical records, as well as those who had resided at altitudes of 2,500 meters above sea level (m.a.s.l.) or higher for more than six consecutive months, were also excluded.

The sample size was estimated using the formula for a single population proportion, utilizing the OpenEpi software (version 3.01). An expected prevalence of abnormal spirometry of 32% was adopted, based on previous studies conducted in occupational populations exposed to similar altitudes in Latin America[14]. Using a 95% confidence level and a 5% margin of error, the minimum required sample size was calculated to be 334 participants. To account for potential exclusions due to invalid spirometry, incomplete data, or ineligibility, an additional 20% was added. Consequently, the final sample size was set at 400 participants, selected by simple random sampling.

### Study Variables

The primary outcome was spirometric abnormality, defined as the presence of any non-normal ventilatory pattern (obstructive, restrictive, or mixed), classified according to ATS/ERS guidelines. Independent variables included age (categorized as <40, 40–50, 50–60, and >60 years), sex, body mass index (BMI), Charlson Comorbidity Index (CCI), job category, duration of employment, prior exposure to harmful agents, severity of COVID-19, and years of intermittent exposure to high altitude.

BMI was categorized according to World Health Organization (WHO) criteria: underweight (<18.5 kg/m²), normal (18.5–24.9 kg/m²), overweight (25.0–29.9 kg/m²), and obesity (≥30 kg/m²). The CCI was classified into no comorbidities (0 points), Low burden (1-2 points), and high burden (≥3 points) comorbidity burden[15]. Occupation was categorized as white-collar (administrative roles) or blue-collar (technical, operational, supervisory, and health, safety, and environment personnel). Employment duration and high-altitude exposure were categorized into three ranges: 3–5 years, 5–7 years, and ≥7 years. COVID-19 severity was classified as mild (outpatient management), moderate (hospitalization), or severe (intensive care unit admission).

### Spirometry Procedures

Spirometry was conducted in Cusco, Peru, at an altitude of 3,400 meters above sea level, using an AMIR Espiro® spirometer, with calibration performed every six months according to the manufacturer’s specifications. The procedures adhered to the 2019 ATS/ERS standards for acceptability, reproducibility, and interpretation. At least three acceptable and two reproducible forced maneuvers were required, each with a forced expiratory time of at least six seconds. The measured parameters included forced vital capacity (FVC), forced expiratory volume in one second (FEV₁), and the FEV₁/FVC ratio.[16]

Ventilatory patterns were interpreted according to ATS/ERS 2021 standards and reference values [17]. The following cut-off values were used:

- Obstructive pattern: FEV₁/FVC < 0.70
- Restrictive pattern: FVC < 80% of predicted with FEV₁/FVC ≥ 0.70
- Mixed pattern: FEV₁/FVC < 0.70 and FVC < 80% of predicted

A spirometry was considered abnormal if it met criteria for any of the above patterns. It was considered normal if FEV₁, FVC, and FEV₁/FVC were all within normal predicted values (i.e., FEV₁ and FVC ≥80% predicted, and FEV₁/FVC ≥0.70).

Only tests rated A or B in quality were accepted. Grade A spirometries fully complied with all technical criteria; grade B tests had minimal deviations that did not compromise interpretation.

### Data Analysis

The data were entered into Microsoft Excel 365 and subsequently analyzed using Stata version 18 (StataCorp LLC, College Station, TX, USA). Data access and analysis were conducted between 1 November 2024 and 30 March 2025. Categorical variables were summarized as absolute and relative frequencies, while continuous variables were described using means and standard deviations. Bivariate analyses were performed using the chi-square or Fisher’s exact test, as appropriate.

For multivariable analysis, a generalized linear model (GLM) was fitted using a Poisson distribution, log link function, and robust variance to estimate adjusted prevalence ratios (aPRs) and their 95% confidence intervals (CIs). Variables with p-values <0.20 in bivariate analysis, along with those of epidemiological relevance (such as age and sex), were included in the model. Collinearity among covariates was evaluated using the variance inflation factor (VIF), ensuring acceptable multicollinearity levels (VIF < 5).

A forest plot was created to display the adjusted prevalence ratios (PRs) with their 95% confidence intervals, using the coefplot command. This visualization summarized the strength of association between covariates and abnormal spirometry.

### Ethical Considerations

This study was conducted in accordance with the ethical principles outlined in the Declaration of Helsinki and national regulations for biomedical research. Approval was granted by the Institutional Committee of Ethics in Research (CIEI) of the Universidad Científica del Sur, under official document CONSTANCIA N°670-CIEI-CIENTÍFICA-2024. The research protocol was registered with the code POS-60-2024-00841. As this study involved secondary analysis of anonymized data collected during routine occupational health evaluations, individual informed consent was not required. However, all personally identifiable information was protected to ensure strict confidentiality.

## Results

### Participant Selection Process

In 2024, a total of 25,423 workers were registered at the occupational health center. Of these, 21,301 were excluded due to either the absence of a confirmed history of COVID-19 or lack of a normal spirometry record from the year 2020. An additional 2,103 individuals were excluded because of invalid post-COVID spirometry results, absence of intermittent high-altitude exposure, or incomplete occupational or clinical records. This resulted in a pool of 2,109 eligible participants, from which a simple random sample of 400 individuals was selected for the study.

### Participant Characteristics

The mean age was 42.5 years (SD ±7.8), with the majority being male (53%). In terms of nutritional status, 46% of the participants were classified as overweight, and 20.5% had a BMI of 30 or higher, meeting criteria for obesity. Clinically, 21.5% of workers had a Charlson Comorbidity Index ≥2, indicating the presence of multiple chronic comorbidities such as hypertension, type 2 diabetes, or cardiovascular disease. These conditions were self-reported and confirmed through clinical records from the occupational health unit. (Table 1)

**Table 1.** Characteristics of the Study Population.

| Category | Frequency<br>N (%) |
| --- | --- |
| **Age** |  |
| < 40 years | 135 (33.8) |
| 40 – 49 years | 100 (25.0) |
| 50 – 59 years | 115 (28.7) |
| 60 – 65 years | 50 (12.5) |
| X ± SD | 47 ± 11.7 |
| **Sex** |  |
| Male | 212 (53.0) |
| Female | 188 (47.0) |
| **Body Mass Index (BMI)** |  |
| Normal (< 25 kg/m<sup>2</sup>) | 134 (33.5) |
| Overweight (25.0–29.9 kg/m<sup>2</sup>) | 184 (46.0) |
| Obesity (≥ 30 kg/m<sup>2</sup>) | 82 (20.5) |
| X ± SD | 26.8 ± 4.0 |
| **Charlson Comorbidity Index** |  |
| No comorbidities | 189 (47.3) |
| Low burden (Charlson Index 1 or 2) | 125 (31.2) |
| High burden (Charlson Index ≥3) | 86 (21.5) |
| **COVID-19 Severity** |  |
| Outpatient management | 241 (60.3) |
| General hospitalization | 124 (31.0) |
| Intensive care | 35 (8.7) |
| **Occupation** |  |
| Administrative | 69 (17.2) |
| Supervisor | 66 (16.5) |
| Environmental Health and Safety | 127 (31.8) |
| Technician | 65 (16.3) |
| Operator | 73 (18.3) |
| **Duration of Employment in Current Company** |  |
| 3 to 5 years | 144 (36.0) |
| 5 to 7 years | 123 (30.7) |
| More than 7 years | 133 (33.2) |
| **Previous Occupational Exposure to Respiratory Irritants** |  |
| No | 186 (46.5) |
| 1 to 3 years | 87 (21.7) |
| 3 to 5 years | 88 (22.0) |
| More than 5 years | 39 (9.7) |
| **Intermittent High-Altitude Exposure** |  |
| Less than 3 years | 165 (41.3) |
| 4 to 6 years | 120 (30.0) |
| 7 or more years | 115 (28.8) |

### COVID-19 Severity and Occupational Exposure

Regarding the clinical course of COVID-19, 60.3% of participants had mild disease managed on an outpatient basis, 31.0% experienced moderate disease requiring hospitalization without intensive care, and 8.7% developed severe disease, defined as ICU admission or the need for supplemental oxygen therapy.

Workers without current exposure to respiratory irritants were included. White-collar (administrative) workers accounted for 17% of the sample. The remaining participants were classified as blue-collar workers. Among them, highly specialized personnel—such as supervisors, technicians, and those responsible for health, safety, and environmental (HSE) areas—represented 64.6%, while low-specialization blue-collar workers, such as operators, accounted for 18.3%. In terms of occupational exposure, 63.4% of the workers have been employed by the company for five years or more and 58% of workers had accumulated more than four years of intermittent high-altitude exposure. Additionally, 9.7% had worked for more than five years in environments with significant exposure to inhaled occupational agents such as silica dust, diesel fumes, or metal welding gases. **(Table 1)**

### Spirometric Findings

Post-pandemic spirometry revealed that 72.2% (n = 289) of the workers had some form of ventilatory abnormality, despite having normal pre-pandemic spirometry in 2020. The most common pattern was mixed (36.5%), followed by restrictive (24.5%) and obstructive (11.3%). According to spirometric classification, 46.2% of participants had an FEV₁ value greater than 70% of the predicted value, while 31.2% had values between 35–49%, and 2.5% had severely reduced FEV₁ (<35%). A total of 38.8% of participants presented with a forced vital capacity (FVC) <80% of the predicted value. Regarding the Tiffeneau Index (FEV₁/FVC), 51.5% had values below 70%, consistent with an obstructive ventilatory pattern, while 48.5% had preserved ratios.**. (Table 2)**

**Table 2.** Spirometric Characteristics and Prevalence of Respiratory Patterns.

| Variable | n | Prevalence (95% CI) |
| --- | --- | --- |
| **FEV<sub>1</sub> Classification (mL)** |  |  |
| >70 | 185 | 46.2% (41.2 – 51.2) |
| 60–69 | 20 | 5.0% (3.1 – 7.6) |
| 50–59 | 60 | 15.0% (11.6 – 18.8) |
| 35–49 | 125 | 31.2% (26.7 – 36.0) |
| <35 | 10 | 2.5% (1.2 – 4.5) |
| **FVC (% of predicted value)** |  |  |
| <80% | 155 | 38.8% (33.9 – 43.7) |
| ≥80% | 276 | 69.0% (64.2 – 73.5) |
| **Tiffeneau Index (FEV<sub>1</sub>/FVC, %)** |  |  |
| <70% | 206 | 51.5% (46.4 – 56.5) |
| ≥70% | 194 | 48.5% (43.5 – 53.5) |
| **Spirometric Alteration** |  |  |
| Normal | 111 | 27.8% (23.4 – 32.4) |
| Abnormal | 289 | 72.2% (67.5 – 76.5) |
| – Restrictive/obstructive pattern | 162 | 40.5% (35.6 – 45.4) |
| – Obstructive pattern only | 83 | 20.8% (16.9 – 25.1) |
| – Restrictive pattern only | 44 | 11.0% (8.1 – 14.5) |

FEV<sub>1</sub> = Forced Expiratory Volume in the first second; FVC = Forced Vital Capacity; 95% CI = 95% Confidence Interval.

### Bivariate and Multivariate Analysis

In bivariate analysis, abnormal spirometry was significantly associated with obesity (p<0.001), Charlson Index ≥2 (p=0.003), moderate-to-severe COVID-19 (p<0.001), high-altitude exposure ≥7 years (p=0.002), and inhalant exposure ≥5 years (p=0.001). **(Table 3)**

**Table 3.** Prevalence of spirometric abnormalities according to demographic, clinical, and occupational characteristics.

| Variable | Abnormal Spirometry | Normal Spirometry | p-Value |
| --- | --- | --- | --- |
| **Sex** |  |  |  |
| Female | 159 (75.0) | 53 (25.0) | 0.192 a |
| Male | 130 (69.1) | 58 (30.9) |  |
| **Age** |  |  |  |
| < 40 years | 102 (75.6) | 33 (24.4) | 0.213 a |
| 40–49 years | 71 (71.0) | 29 (29.0) |  |
| 50–59 years | 76 (68.0) | 39 (33.9) |  |
| 60–65 years | 40 (80.0) | 10 (20.0) |  |
| **Body Mass Index (BMI)** |  |  |  |
| Normal (<25 kg/m<sup>2</sup>) | 92 (68.7) | 42 (31.3) | < 0.001a |
| Overweight (25–29.9 kg/m<sup>2</sup>) | 115 (62.5) | 69 (37.5) |  |
| Obesity (≥30 kg/m<sup>2</sup>) | 82 (100.0) | 0 (0.0) |  |
| **Charlson Comorbidity Index** |  |  |  |
| No comorbidities | 119 (63.0) | 70 (37.0) | < 0.001 a |
| Low burden (Charlson Index 1 or 2) | 84 (67.2) | 41 (32.8) |  |
| High burden (Charlson Index ≥3) | 86 (100.0) | 0 (0.0) |  |
| **COVID-19 Severity** |  |  |  |
| Outpatient management | 172 (71.3) | 69 (28.6) | < 0.001 a |
| General hospitalization | 82 (66.1) | 42 (33.9) |  |
| Intensive care | 35 (100.0) | 0 (0.0) |  |
| **Occupation** |  |  |  |
| Administrative | 48 (69.6) | 21 (30.4) | 0.9280 a |
| Supervisor | 44 (68.7) | 20 (31.3) |  |
| Environmental, health and safety | 101 (73.2) | 37 (26.8) |  |
| Technician | 47 (74.6) | 16 (25.4) |  |
| Operator | 49 (74.2) | 17 (25.7) |  |
| **Duration of employment** |  |  |  |
| 3–4.9 years | 106 (73.6) | 38 (26.4) | 0.762 b |
| 5–7.9 years | 90 (73.2) | 33 (26.8) |  |
| ≥7 years | 93 (69.9) | 40 (30.1) |  |
| **Previous Occupational Exposure to Respiratory Irritants** |  |  |  |
| None | 121 (65.1) | 65 (34.9) | < 0.001 a |
| 1–2.9 years | 65 (74.7) | 22 (25.3) |  |
| 3–4.9 years | 64 (72.7) | 24 (27.3) |  |
| ≥5 years | 39 (100.0) | 0 (0.0) |  |
| **Intermittent High-Altitude Exposure** |  |  |  |
| <4 years | 94 (56.9) | 71 (43.1) | < 0.001 a |
| 4–6 years | 80 (66.7) | 40 (33.3) |  |
| ≥7 years | 115 (100.0) | 0 (0.0) |  |

<sup>a</sup>Chi cuadrado <sup>b</sup>U Mann-Whitney

The frequency of spirometric pattern abnormalities across clinical and occupational categories is presented in **S1 Table,** while descriptive statistics of spirometric indices (FEV₁, FVC, and FEV₁/FVC ratio) by demographic, clinical, and occupational characteristics are summarized in **S2 Table**.

In multivariate Poisson regression, independent predictors of spirometric alteration were obesity (aPR: 1.49; 95% CI: 1.33–1.68), Charlson Index ≥2 (aPR: 1.49; 95% CI: 1.33–1.68), severe COVID-19 (aPR: 1.65; 95% CI: 1.41–1.91), high-altitude exposure ≥7 years (aPR: 1.81; 95% CI: 1.60–2.05), and inhalant exposure ≥5 years (aPR: 1.64; 95% CI: 1.43–1.89), all with p<0.01. (**Table 4 and Fig 2)**

**Fig 1.**
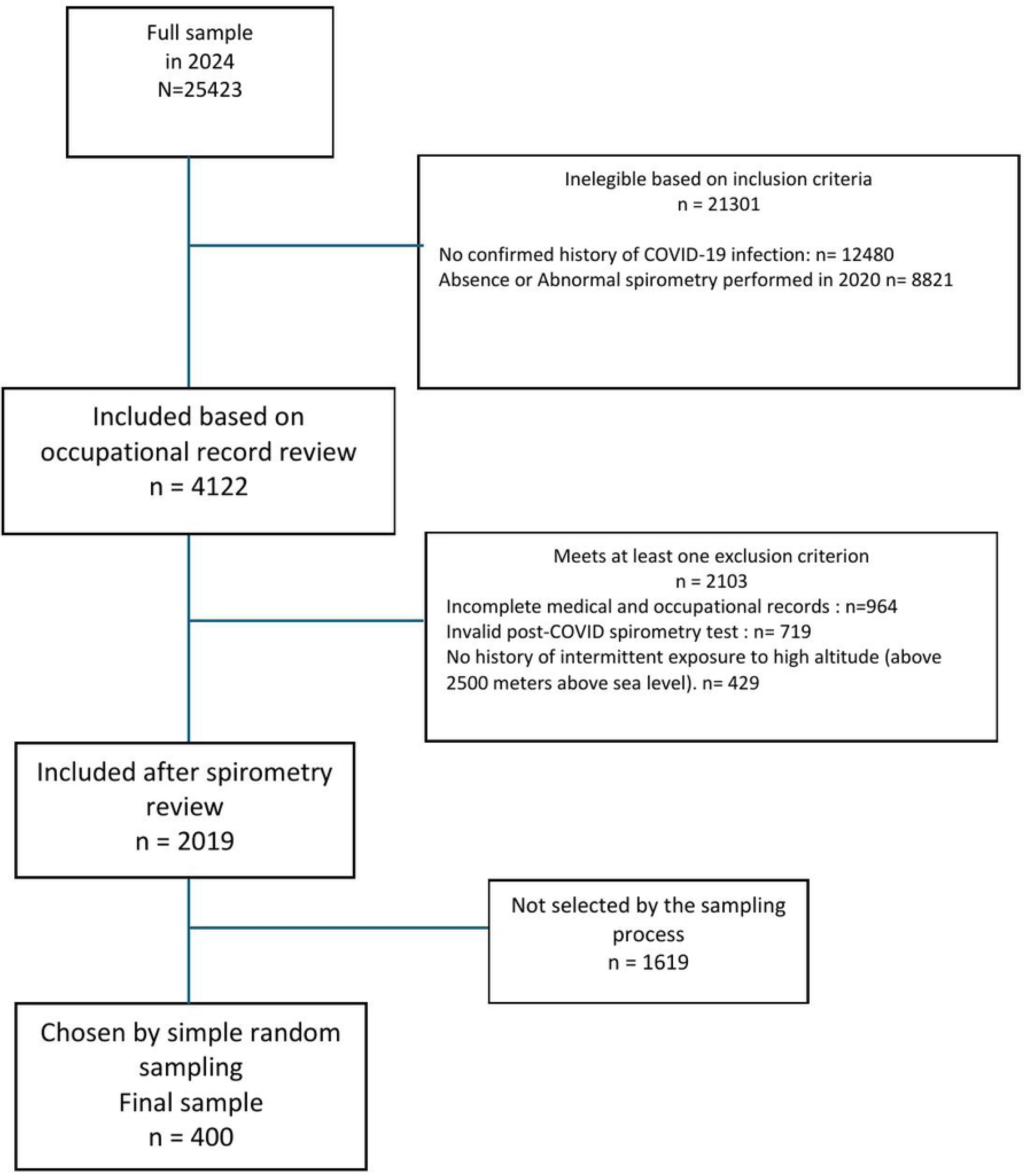
Flowchart of Study Population Selection.

**Fig 2.**
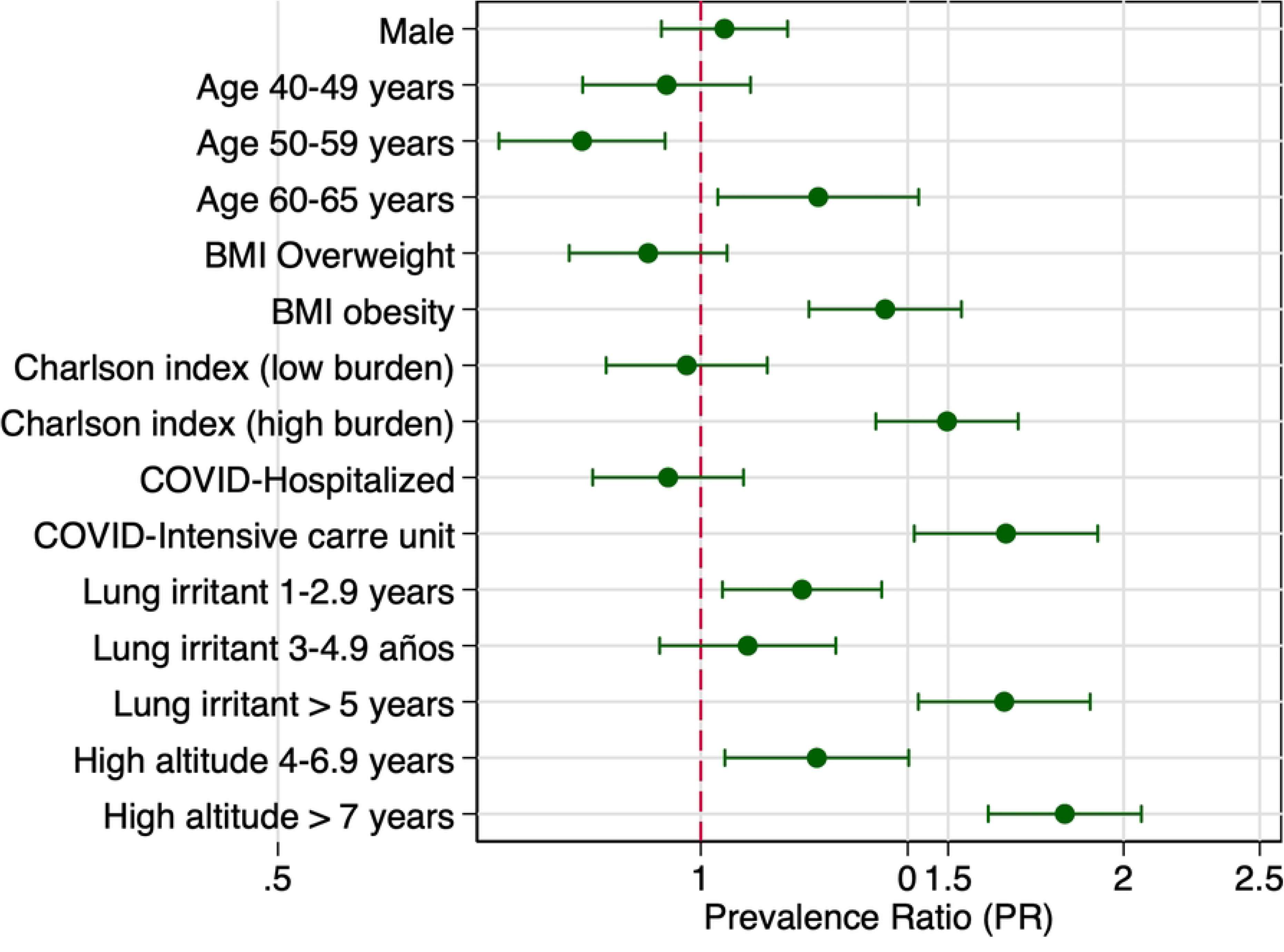
Forest plot of adjusted prevalence ratios for abnormal spirometry.

**Table 4.** Prevalence ratios for abnormal spirometry according to demographic and clinical factors in workers with intermittent high-altitude exposure.

| Variable | Crude PR (95% CI) | p-value | Adjusted PR (95% CI) | p-value |
| --- | --- | --- | --- | --- |
| **Sex** |  |  |  |  |
| Female | Reference | — | Reference | — |
| Male | 1.08 (0.95 – 1.22) | 0.197 | 1.03 (0.93 – 1.15) | 0.462 |
| **Age** |  |  |  |  |
| < 40 years | Reference | — | Reference | — |
| 40–49 years | 0.93 (0.80 – 1.10) | 0.440 | 0.94 (0.82 – 1.09) | 0.425 |
| 50–59 years | 0.87 (0.74 – 1.03) | 0.106 | 0.82 (0.71 – 0.94) | 0.718 |
| 60–65 years | 1.05 (0.89 – 1.25) | 0.507 | 1.21 (1.02 – 1.42) | 0.022 |
| **Body Mass Index (BMI)** |  |  |  |  |
| Normal (<25 kg/m<sup>2</sup>) | Reference | — | Reference | — |
| Overweight (25–29.9 kg/m<sup>2</sup>) | 0.91 (0.77 – 1.06) | 0.250 | 0.92 (0.81 – 1.04) | 0.191 |
| Obesity (≥30 kg/m<sup>2</sup>) | 1.45 (1.29 – 1.63) | <0.001 | 1.35 (1.19 – 1.53) | <0.001 |
| **Charlson Comorbidity Index** |  |  |  |  |
| No comorbidities | Reference | — | Reference | — |
| Low burden (Charlson Index 1 or 2) | 1.06 (0.91 – 1.26) | 0.437 | 0.98 (0.86 – 1.11) | 0.733 |
| High burden (Charlson Index ≥3) | 1.59 (1.42 – 1.77) | <0.001 | 1.49 (1.33 – 1.68) | <0.001 |
| **COVID-19 Severity** |  |  |  |  |
| Outpatient management | Reference | — | Reference | — |
| General hospitalization | 0.92 (0.79 – 1.08) | 0.317 | 0.95 (0.84 – 1.07) | 0.395 |
| Intensive care | 1.40 (1.29 – 1.51) | <0.001 | 1.65 (1.41 – 1.91) | <0.001 |
| **Occupation** |  |  |  |  |
| Administrative | Reference |  |  |  |
| Supervisor | 0.99 (0.79 – 1.24) | 0.919 |  |  |
| Environmental, health and safety | 1.05 (0.87 – 1.26) | 0.593 |  |  |
| Technician | 1.07 (0.87 – 1.33) | 0.519 |  |  |
| Operator | 1.07 (0.86 – 1.31) | 0.546 |  |  |
| **Duration of employment** |  |  |  |  |
| 3–4.9 years | Reference | — |  |  |
| 5–6.9 years | 0.99 (0.86 – 1.15) | 0.935 |  |  |
| ≥7 years | 0.95 (0.82 – 1.11) | 0.498 |  |  |
| **Previous Occupational Exposure to Respiratory Irritants** |  |  |  |  |
| None | Reference | — | Reference | — |
| 1–2.9 years | 1.14 (0.97 – 1.35) | 0.093 | 1.18 (1.04 – 1.35) | 0.013 |
| 3–4.9 years | 1.12 (0.94 – 1.32) | 0.188 | 1.08 (0.93 – 1.25) | 0.296 |
| ≥5 years | 1.53 (1.38 – 1.71) | <0.001 | 1.64 (1.43 – 1.89) | <0.001 |
| **Intermittent High-Altitude Exposure** |  |  |  |  |
| <4 years | Reference | — | Reference | — |
| 4–6.9 years | 1.17 (0.97 – 1.40) | 0.093 | 1.21 (1.04 – 1.40) | 0.013 |
| ≥7 years | 1.75 (1.53 – 2.00) | <0.001 | 1.81 (1.60 – 2.05) | <0.001 |

PR = Prevalence Ratio; CI = Confidence Interval

## Discussion

### Main Findings

This study found that 72.3% of high-altitude workers with a prior history of SARS-CoV-2 infection developed spirometric abnormalities, despite having had normal pulmonary function before the pandemic. The most frequent ventilatory pattern was mixed (36.5%), followed by restrictive (24.5%) and obstructive (11.3%) types. The multivariate analysis identified five independent predictors of spirometric alteration: obesity, a Charlson Comorbidity Index ≥2, severe COVID-19, cumulative high-altitude exposure of seven or more years, and inhalant exposure for five or more years. These results suggest a significant burden of post-COVID-19 pulmonary dysfunction in a working population exposed to extreme environmental conditions such as intermittent hypobaric hypoxia and occupational inhalants and reflect a multifactorial interaction between biological vulnerability and environmental stressors.

### Comparison with Other Studies

The prevalence of abnormal spirometry in our study (72.2%) is significantly higher than that reported in other post-COVID-19 cohorts with different altitude exposures. For example, a cross-sectional study in Malaysia, conducted at sea level, found a 46.8% prevalence of abnormal spirometry, mainly characterized by restrictive and Preserved Ratio Impaired Spirometry (PRISm) patterns, with only 1.0% showing obstructive alterations [18]. PRISm is defined as reduced FEV₁ (< 80% predicted) with preserved FEV₁/FVC ratio (≥ 0.70), indicating diminished lung function without classical obstruction. A global meta-analysis by Torres-Castro et al. reported 7% prevalence of obstructive spirometry, 15% restrictive pattern, and 39% impaired diffusing capacity (DLCO), primarily among individuals at sea level recovering from varying severities of COVID-19.[19] In contrast, a multicenter study conducted in Latin America (FIRCOV study), which included populations permanently residing at moderate and high altitudes, found a 32% prevalence of restrictive spirometry pattern in post-COVID-19 individuals, along with a 43.7% prevalence of reduced diffusing capacity (DLCO)[20]. Notably, our study focused on workers with intermittent exposure to high altitude, who had normal spirometry prior to COVID-19 and were assessed months after recovery. This occupational exposure pattern—characterized by repeated cycles of hypobaric hypoxia—may exacerbate pulmonary vulnerability, potentially explaining the higher prevalence of mixed restrictive/obstructive patterns observed. These findings highlight the need to consider altitude exposure type (permanent vs. intermittent) as a potential modifier of post-COVID pulmonary outcomes.

In several studies conducted primarily at sea level, post-COVID-19 abnormal spirometry has been associated with a variety of factors. From a demographic standpoint, a higher prevalence of alterations has been observed in individuals over 50 years of age and in males, as reported in studies from Bangladesh and India[21,22]. Regarding clinical factors, disease severity, prolonged hospitalization, and the use of oxygen therapy or non-invasive ventilation have been linked to restrictive and mixed spirometric patterns[23,24]. Additionally, comorbidities such as hypertension, diabetes, cardiovascular disease, and smoking history have been identified as significant predictors [21,25]. In terms of imaging findings, studies from Thailand and Europe have shown that the presence of consolidations, fibrosis, or ground-glass opacities on chest imaging is significantly associated with spirometric impairment, even in patients with seemingly preserved lung function[24,26]. Unlike these studies, our research was conducted in a setting of intermittent high-altitude exposure, contributing valuable evidence from a less-explored environment..

### Interpretation According to Occupational and Clinical Factors

The predominance of mixed ventilatory patterns suggests that the pathophysiological mechanisms in this cohort involve both airway and interstitial involvement.[27] Persistent low-grade inflammation or fibrosis due to SARS-CoV-2 may interact synergistically with pre-existing occupational exposures to damage alveolar-capillary units and small airways.[28] Exposure to high concentrations of silica particles and diesel exhaust has been shown to induce oxidative stress, inflammatory cytokine release, and airway remodeling.[29] Moreover, hypobaric hypoxia at high altitudes can cause chronic pulmonary hypertension, reduced diffusing capacity, and impaired alveolar regeneration.[30]

Although no studies have directly evaluated how age, body mass index (BMI), prior COVID-19 severity, comorbidity burden, duration of intermittent high-altitude exposure, and previous exposure to respiratory irritants affect spirometric outcomes among currently healthy workers intermittently exposed to high altitude, evidence from related clinical and occupational research supports plausible inferences. In general populations, older age and elevated BMI are independently associated with restrictive and mixed ventilatory patterns, likely due to altered chest wall mechanics and reduced thoracic compliance with aging [31,32]. Patients recovering from moderate to critical COVID-19 frequently present persistent spirometric abnormalities— approximately 28% restrictive and 17% PRISm—especially in those with radiographic lung changes, desaturation, or ICU admission[33]. A higher number of pre-existing comorbidities (e.g., cardiovascular disease, diabetes, COPD) further increases the risk of long-term pulmonary impairment post-COVID-19.[34] Longer cumulative exposure to high-altitude work (≥7 years) was independently associated with altered spirometry. Previous studies in Andean miners have documented chronic mountain sickness and hypoxemia-related structural lung changes even in the absence of infection[35,36]. It is plausible that chronic hypoxia created a “primed” lung environment, less resilient to the inflammatory insult of SARS-CoV-2. Similarly, five or more years of inhalant exposure was associated with a 64% higher prevalence of spirometric alterations.[37] Although biological sex influences lung anatomy—males generally have ∼12% larger lung volumes and wider airways—even without percent-predicted normalization, our study found no sex-based differences in spirometric outcomes. This likely reflects a relatively homogeneous cohort of healthy workers with similar age, BMI, comorbidity profiles, and exposure history, where anthropometric variation was minimal. Additionally, disease-specific sex effects such as higher PRISm prevalence in females often emerge only in clinically diverse or larger populations.[38]

### Public Health and Occupational Medicine Implications

These findings have significant implications for occupational health surveillance and policy in high-altitude settings. First, the high prevalence of subclinical spirometric abnormalities, even in workers who report no persistent symptoms, challenges current return-to-work protocols that rely predominantly on symptom resolution. Occupational medicine programs should consider incorporating post-COVID spirometric assessments into their functional medical evaluations for workers exposed to extreme environments, such as high altitudes—particularly for those with identified risk factors such as obesity, comorbidities, or a history of moderate to severe COVID-19..[39]

Second, our results support the integration of baseline spirometry into pre-placement evaluations, as it enables future comparison and detection of change over time. The presence of documented normal pre-pandemic lung function in our cohort significantly strengthens causal inference. This reinforces the value of longitudinal data and occupational health records in epidemiological investigations.[40,41]

Third, workplace interventions such as improved ventilation systems, targeted respiratory protective equipment, and scheduled medical surveillance may help mitigate risk in those with known inhalant exposures. Rehabilitation programs tailored for high-altitude reintegration, including graded physical reconditioning and respiratory physiotherapy, may improve outcomes for affected workers.[42]

Finally, considering that Peru has a vast high-Andean occupational territory, it is recommended that the Ministry of Health develop protocols or a guideline on Occupational Exposure to Chronic Intermittent Hypobaria at High Altitude, based on evidence-informed data.

### Strengths and Limitations

This study has several strengths. First, the availability of pre-pandemic spirometry for all included participants enables robust comparison and supports a temporal link between COVID-19 and observed abnormalities. Second, the inclusion of multiple clinically and occupationally relevant variables enhances the external validity of the findings for similar worker populations in the Andean and Himalayan regions. Third, the use of a standardized spirometry protocol following ATS/ERS 2019 guidelines, and reference values adjusted to South American populations, ensures technical accuracy and reproducibility.[16]

However, some limitations must be acknowledged. The cross-sectional design precludes evaluation of the evolution of lung function over time or assessment of reversibility. Diffusion capacity for carbon monoxide (DLCO) and high-resolution computed tomography (HRCT), which could have clarified underlying mechanisms such as fibrosis or vascular abnormalities, were not performed. Similarly, the study was conducted at sea level, and post-exertional desaturation at altitude could not be assessed. The lack of female representation limits generalizability to women in similar occupations, and reliance on occupational health records may introduce reporting bias in exposure assessment.

### Recommendations for Future Research

Longitudinal studies are needed to track the progression or resolution of post-COVID spirometric changes over time, especially in high-risk subgroups. These studies should incorporate comprehensive pulmonary function testing, including DLCO, lung volumes, and imaging. Evaluating cardiopulmonary exercise capacity and hypoxia challenge testing could provide further insight into functional limitations at altitude. Molecular studies exploring inflammatory markers or genetic predisposition to post-viral fibrosis in hypoxic environments would enhance understanding of pathogenesis.

Additionally, future research should examine the effectiveness of tailored interventions, including pulmonary rehabilitation and ergonomic reassignments, in restoring work capacity and preventing further respiratory decline. Health economic evaluations could guide cost-effective implementation of screening and follow-up programs in resource-limited occupational settings.

At the occupational level, each occupational physician should assess the relevance of incorporating functional medical tests that are appropriate to the specific work sector. In addition, as part of health surveillance, occupational physicians should guide workers to attend both occupational and clinical evaluations when pathology is suspected, and correlate the findings to enhance the effectiveness of health surveillance

### Conclusions

In conclusion, our study demonstrates a high prevalence of spirometric abnormalities among high-altitude workers previously infected with COVID-19, particularly those with comorbidities, obesity, severe disease, and long-term occupational exposures. The predominance of mixed ventilatory patterns reflects a complex interplay between viral injury and environmental stressors such as inhalants and hypoxia. These findings support the need for structured respiratory surveillance and preventive interventions in occupational health systems for high-altitude populations. Incorporating spirometry into routine post-COVID-19 evaluations can help detect subclinical impairments and guide safe reintegration into high-risk workplaces.

## Data Availability

All relevant data are within the manuscript and its Supporting Information files.

## Supplementary tables

**S1 Table. Frequency of Spirometric Patterns by Clinical and Occupational Characteristics**

**S2 Table. Spirometric Indices (FEV₁, FVC, and FEV₁/FVC Ratio) by Demographic, Clinical, and Occupational Characteristics"**

